# Single-Cell Analysis Reveals Macrophage-Driven T Cell Dysfunction in Severe COVID-19 Patients

**DOI:** 10.1101/2020.05.23.20100024

**Authors:** Xiaoqing Liu, Airu Zhu, Jiangping He, Zhao Chen, Longqi Liu, Yuanda Xu, Feng Ye, Huijian Feng, Ling Luo, Baomei Cai, Yuanbang Mai, Lihui Lin, Zhenkun Zhuang, Sibei Chen, Junjie Shi, Liyan Wen, Yuanjie Wei, Jianfen Zhuo, Yingying Zhao, Fang Li, Xiaoyu Wei, Dingbin Chen, Xinmei Zhang, Na Zhong, Yaling Huang, He Liu, Jinyong Wang, Xun Xu, Jie Wang, Ruchong Chen, Xinwen Chen, Nanshan Zhong, Jingxian Zhao, Yi-min Li, Jincun Zhao, Jiekai Chen

## Abstract

The vastly spreading COVID-19 pneumonia is caused by SARS-CoV-2. Lymphopenia and cytokine levels are tightly associated with disease severity. However, virus-induced immune dysregulation at cellular and molecular levels remains largely undefined. Here, the leukocytes in the pleural effusion, sputum, and peripheral blood biopsies from severe and mild patients were analyzed at single-cell resolution. Drastic T cell hyperactivation accompanying elevated T cell exhaustion was observed, predominantly in pleural effusion. The mechanistic investigation identified a group of CD14^+^ monocytes and macrophages highly expressing *CD163* and *MRC1* in the biopsies from severe patients, suggesting M2 macrophage polarization. These M2-like cells exhibited up-regulated *IL10, CCL18, APOE, CSF1* (M-CSF), and *CCL2* signaling pathways. Further, SARS-CoV-2-specific T cells were observed in pleural effusion earlier than in peripheral blood. Together, our results suggest that severe SARS-CoV-2 infection causes immune dysregulation by inducing M2 polarization and subsequent T cell exhaustion. This study improves our understanding of COVID-19 pathogenesis.

## INTRODUCTION

Coronavirus Disease 2019 (COVID-19) caused by a novel coronavirus, severe acute respiratory syndrome coronavirus 2 (SARS-CoV-2), was first reported in December 2019 in Wuhan, Hubei Province, China. As of April 24, 2020, a total of 2,626,321 cases with 181,938 deaths have been confirmed globally, as reported by the World Health Organization (WHO). This pandemic has quickly become a unprecedent major public emergency of global concerns (Chan et al., 2020; Chen et al., 2020; Huang et al., 2020; Li et al., 2020b).

Most COVID-19 patients experience mild symptoms. However, a significant proportion of the patients develop severe pneumonia and require ventilator-assisted breathing (Guan et al., 2020; Huang et al., 2020; Wang et al., 2020). Patients with severe diseases have significantly higher levels of inflammatory response in their plasma compared to patients with mild disease, indicating dysregulation of immune responses (Huang et al., 2020). A recent autopsy study of COVID-19 patients has also revealed macrophage infiltration and excess production of mucus in the infected lungs, especially in the damaged small airways and alveoli (Liu et al., 2020b).

Upon respiratory CoV infection, properly regulated immune response is essential to control and eliminate the virus as well as to improve clinical outcome (Braciale et al., 2012), whereas maladjusted immune responses may result in immunopathology and impaired pulmonary gas exchange (Chen and Subbarao, 2007; de Wit et al., 2016; Li et al., 2020a). Increased levels of serum proinflammatory cytokines (e.g., IL-1β, IL-6, IL-12, IFN-γ, IP-10, and MCP-1) are associated with pulmonary inflammation and extensive lung damage in SARS patients (Wong et al., 2004). Meanwhile, another set of proinflammatory cytokines, including IFN-γ, TNF, IL-15, and IL-17 were induced in Middle East Respiratory Syndrome coronavirus (MERS-CoV) infection (Mahallawi et al., 2018). The disease-specific cytokine profiles indicate that different host immune factors play a role in the pathogenesis of these highly pathogenic CoV infections. A recent study has reported that serum inflammatory cytokine profiles in severe COVID-19 patients is similar to that of SARS-CoV infection, with elevated concentrations of IL-1β, IFN-γ, IP-10, and MCP-1 (Huang et al., 2020). In addition, either cytokine storm-relevant factors such as G-CSF and TNF, or Th2-related cytokines such as IL-4 and inhibitory IL-10, were found with SARS-CoV-2 infection (Huang et al., 2020). These results suggest that overall balance of immune responses may be important in disease progression and host recovery from respiratory CoV infections.

In SARS-CoV infected mice, we have found that exuberant inflammatory responses and lung damage associated with dysregulated cytokine response induce the accumulation of pathogenic inflammatory monocyte-macrophages (IMMs) in the lung. The IMMs not only elevated lung cytokine/chemokine levels but also inhibited SARS-CoV-specific T cell responses in mice, leading to delayed viral clearance and deteriorated clinical outcomes (Channappanavar et al., 2016). In MERS-CoV-infected mice, early type-I interferon treatment (IFN-I) was protective, whereas delayed IFN-I treatment failed to effectively inhibit virus replication, increased infiltration and activation of monocytes and macrophages in the lungs, and enhanced proinflammatory cytokine expression, resulting in fatal pneumonia (Channappanavar et al., 2019). These studies suggest that not only proper cytokine response but also inflammatory monocytes and/or macrophages play crucial roles in the respiratory CoV pathogenesis.

CoV-specific T cells are required for viral clearance and for protection from clinical disease (Channappanavar et al., 2014; Zhao et al., 2010). We have shown in a mouse model that SARS-CoV-specific CD4^+^ T cells in the airway promoted anti-viral innate immune response and viral-specific CD8^+^ T cell response by increasing respiratory dendritic cell migration from the lung to the draining lymph nodes (Zhao et al., 2016). In MERS-CoV infection, the recovery of patients from MERS is also associated with CD8^+^ T cell responses. MERS patients with robust viral-specific CD8^+^ T cell response in peripheral blood mononuclear cell (PBMC) but not viral-specific CD4^+^ T cell response spent a shorter period in the ICU (Zhao et al., 2017). It is likely that T cell responses also play an important role in viral clearance and host recovery from SARS-CoV-2 infection. However, the phenotype and function of T cells, their behavior in the proinflammatory microenvironment, and their interaction with other immune cells have not been elucidated in COVID-19 patients, particularly T cells in human lungs which are most relevant clinically yet difficult to obtain from human patients compared to PBMC.

Here, by integrating advanced single-cell technology and immunological approaches, leukocytes derived from the pleural effusion, sputum, and peripheral blood biopsies of severe and mild COVID-19 patients were analyzed. SARS-CoV-2-specific T cells were detected in pleural fluid mononuclear cells (PFMC) earlier than in PBMC and correlated with viral clearance. Immune dysregulation and T cell exhaustion were observed, together with accumulation of T cell suppressive M2-macrophages in PFMC. Further analyses suggested that severe SARS-CoV-2 infection induced M2-macrophage polarization in the lung that might play a role in driving T cell exhaustion. These findings and mechanistic insights not only improved our understanding of COVID-19 pathogenesis and mechanism for immune dysregulation, but also demonstrated the value of pleural effusion in translational research with implications for disease diagnostics and treatment.

## RESULTS

### scRNA-seq revealed distinct immune cell composition and state in the COVID-19 patient

A 70s old man infected with SARS-CoV-2 (confirmed by real-time PCR) developed severe pneumonia and was admitted to the intensive care unit (ICU) of the First affiliated hospital of Guangzhou Medical University. Invasive ventilator-assisted breathing was instituted at 9 days post symptom onset (d.p.o.). Thymosin (Zadaxin) treatment was started from 10 d.p.o. Pleural effusion was observed at 19 d.p.o. and drainage tube was placed. Pleural fluid (at 20 d.p.o.) and serial peripheral blood were collected (Figure 1A). To characterize the immune responses in humans upon SARS-CoV-2 infection, single-cell RNA-sequencing (scRNA-seq) was performed on the patient’s paired pleural fluid mononuclear cells (PFMC) and peripheral blood mononuclear cells (PBMC) obtained at 20 d.p.o. Passing through rigorous quality-control processes, the transcriptome profiles of 7,587 PFMC and 3,874 PBMC cells were subjected to subsequent analysis (Figures 1B and S1A). A PBMC scRNA-seq dataset (Zheng et al., 2017) composed of 9,707 cells from a healthy individual was repurposed and integrated as a control, denoted H-PBMC. Most cell types were conserved among the PFMC, PBMC, and H-PBMC samples, including CD4^+^/CD8^+^ T cell subsets, B cells, NK cells, monocytes and macrophages, to a much less extent dendritic cells (DC) and plasmacytoid DC (pDC) (Figures 1C-E, S1B). Consistent with the finding of lymphopenia reported in other COVID-19 patients (Diao et al., 2020; Liu et al., 2020a), absolute cell count assay showed that CD8^+^ T cell numbers in the patient’s blood dropped far below the normal range (Figure S1C), with only a slight decrease for CD4^+^ T cells and NK cells, and no significant change for B cells (Figure S1C), resulted in increased CD4^+^/CD8^+^ T cell ratios in both PBMC and PFMC comparing to H-PBMC (Figure S1D). CCR7^+^ naïve CD4^+^ T cells were predominant in the patient’s PBMC, while PFMC contained more activated CD4^+^ T and CD8^+^ T cells by frequency (Figures 1C,1D, S1D and S1E). The percentage of FOXP3^+^CD25^+^ Treg and CD25 expression were higher in PFMC, and a group of IL10+ T cells was exclusive in the patient’s PFMC (Figures 1C, 1D and S1F and S1G). Conventional cell lineage markers and flow cytometry analysis further validated the clustering and annotation results (Figures 1D, 1E and S1B). These data reflected the changes in immune cell composition in pulmonary and peripheral in response to viral infection. Of note, the expression of SARS-CoV-2 entry receptor angiotensin-converting enzyme II (ACE2) was also examined in scRNA-seq (Zhou et al., 2020). Minimal ACE2 expression was found in all immune cell types (Figure S1H).

**Figure 1.**
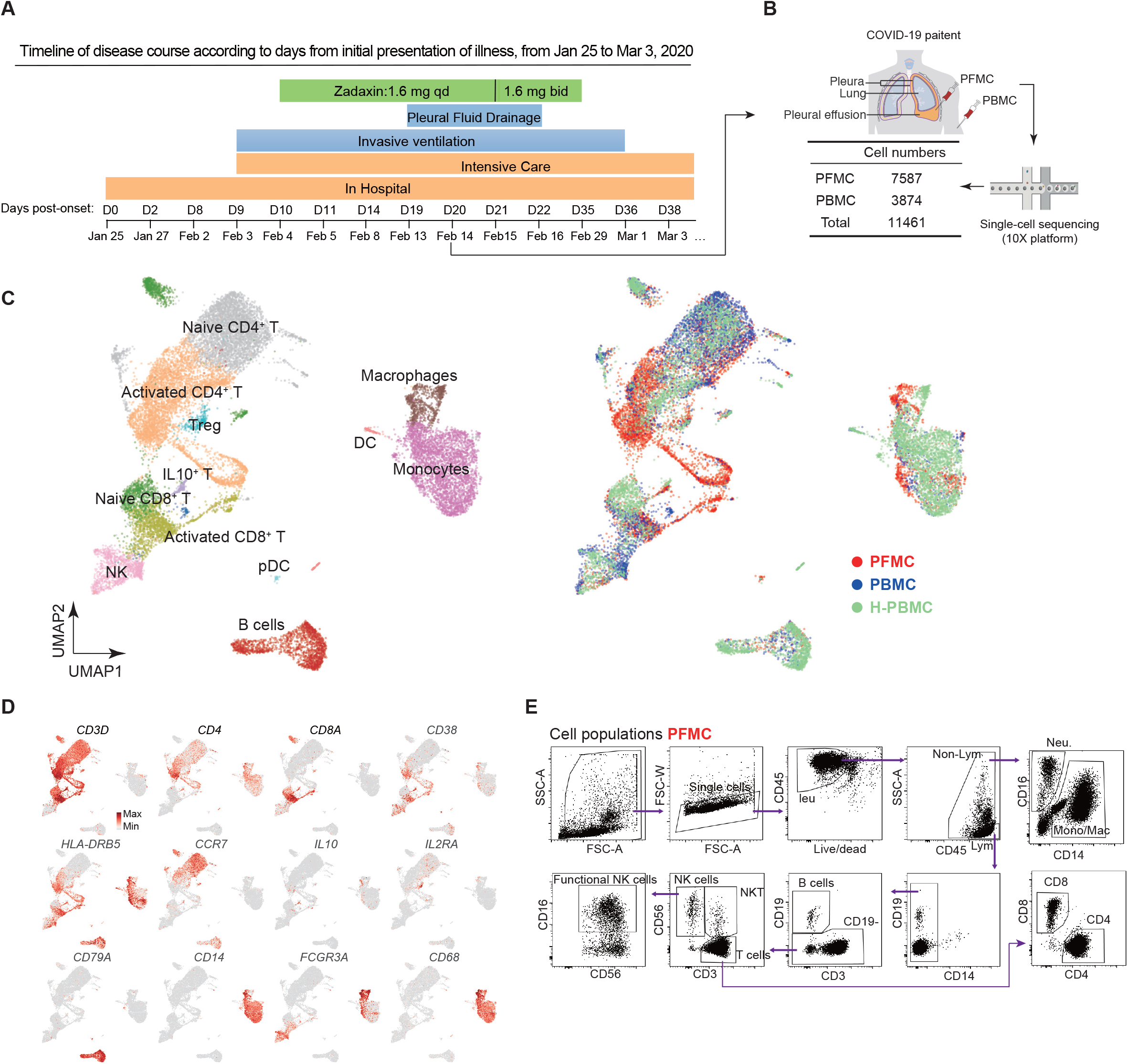
A Comprehensive Survey of Single Cell Reveals Unique Immune State for the COVID-19 Patient. (A) Schematic diagram showing the timeline of disease course of the patient. (B) Schematic diagram showing the isolation of pleural fluid mononuclear cells (PFMC) and peripheral blood mononuclear cells (PBMC) from the COVID-19 patient. Single-cell sequencing was taken by 10x platform. (C) (Left) UMAP plots visualizing single-cell RNA-seq data of 7,587 PFMC single cells and 3874 PBMC single cells from the COVID-19 patient and 9,707 PBMC single cells from a healthy donor (H-PBMC). (Right) Differences in cellular composition among PFMC, PBMC and H-PBMC. (D) UMAP plots showing the expression of maker genes for particular cell types. Gene expression levels are indicated by shades of red. (E) Flow cytometry dot plots showing the identification of various leukocyte subpopulations in the PFMC.

### Significant T cell hyperactivation and exhaustion were detected in the COVID-19 patient

To investigate signaling transduction regulation associated with SARS-CoV-2 infection in human, ligand-receptor interaction analysis among seven major cell types (i.e., naive/activated CD4^+^/CD8^+^ T cell, NK cell, macrophage, and monocyte) was performed. More interactions were instigated than abolished upon infection (Figures 2A and S2A). Explicitly, CCR5-associated pathways involving chemokines CCL3, CCL4, CCL5 and CXCR4 were induced in the activated CD8^+^ T cells in the patient’s PFMC and PBMC, prominently in PFMC, suggesting CD8^+^ T cell hyperactivation (Figures 2A-C and S2B) (Contento et al., 2008; Dairaghi et al., 1998; Honey, 2006; Trifilo et al., 2003). The CCL2-CCR4 pathway was initiated in the activated CD4^+^ population accompanied by an elevated fraction of activated CD4^+^ cells which was observed exclusively in PFMC (Figures 2B-2D and S2B). This was further confirmed by flow cytometry analysis showing a higher proportion of CD4^+^ T cells expressed CCR4 in PFMC than in PBMC (Fig. S2C). CCR4 is a key chemokine receptor guiding T cell to the lung (D’Ambrosio et al., 2001), suggesting CCR4-expressing T cells would be preferentially recruited to the infected site. We also observed mild and strong induction of activated T-cell (CD38^+^HLA-DR^+^) population in PBMC and PFMC, respectively (Figures 2D). Of note, the majority of these activated T cells highly expressed PD-1 (Figure 2E), suggesting T-cell exhaustion upon activation. The percentage of CD38^+^HLA-DR^+^PD1^+^ triple-positive CD4^+^ and CD8^+^ cells was significantly higher in PFMC than in the patient’s PBMC indicating massive T-cell exhaustion in PFMC where could more closely reflect the status of T cells in the lung local inflammatory microenvironment (Figure 2F). Consistently, T cell exhaustion markers, *PDCD1* (PD1), *LAG3, HAVCR2* (TIM3), and *PRDM1* had enhanced expression in PFMC in scRNA-seq analysis (Figures 2G and S2D; Table S1). Further, activated T cells in the patient’s PFMC and PBMC also lost IL-2, IL-7R and TNF expressions (Figure S2A) which are critical for T cell proliferation, survival, as well as signs for early T cell exhaustion (Wherry et al., 2007; Yi et al., 2010). NK and B cell responses to the infection were also investigated. NK/B cell activation pathway genes including *CD160, KLRK1* and *CD22* (Clark and Lane, 1991; Haas et al., 2018; Le Bouteiller et al., 2011; Orr and Lanier, 2010) were found to be significantly down-regulated in patient’s PFMC and PBMC (Figures S2D and S2E; Table S1). Taken together, these results demonstrated that T cells in COVID-19 patients underwent significant hyperactivation and exhaustion upon prolonged SARS-CoV-2 infection, especially in the lung which could contribute to the delayed viral clearance.

**Figure 2.**
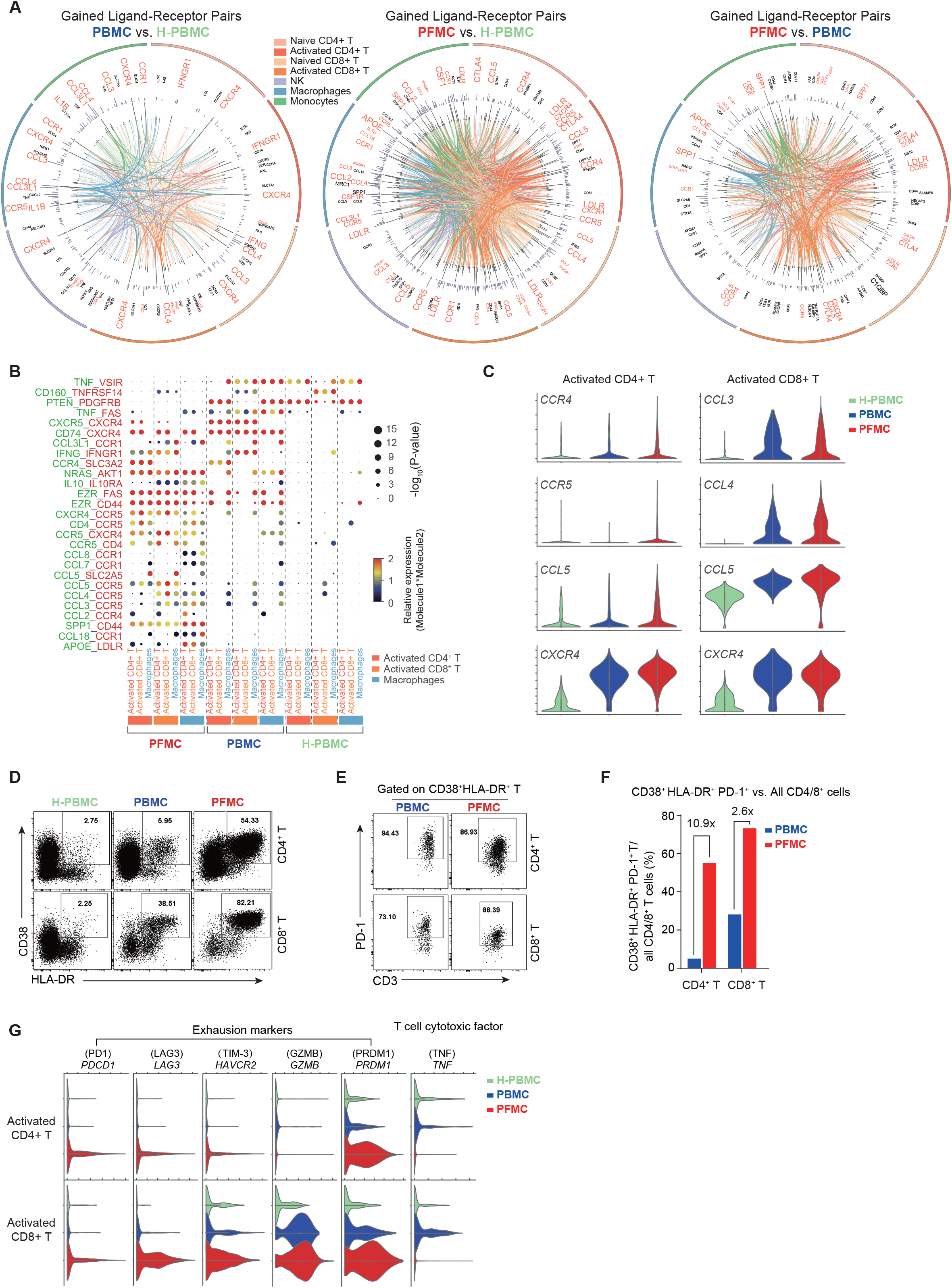
Multiple Regulatory Immune Responses for COVID-19 patient. (A) Circos plots showing the ligand-receptor interactions across different cell types. The outermost ring represents different cell types according to the color bar. The innermost color lines represent ligand-receptor contacts in indicated cell types, the line thickness represents the contact strength. The adjacent black ring shows ligand-receptor interaction strength changes in between indicated samples. The next black ring represents the expression changes of indicated genes in the indicated cell types. (B) Overview of ligand-receptor interactions of selected cytokines, P-value indicated by the dot size. The product of the means of average expression level of molecule 1 (ligand, green) and molecule 2 (receptor, red) are indicated by color. (C) Violin plots showing the expression changes of selected genes in the indicated cell types. (D) Flow cytometry dot plots showing the expression of CD38 and HLA-DR by CD4^+^ and CD8^+^ T cells. (E) Flow cytometry dot plots showing PD-1 expression by CD38^+^HLA-DR^+^ activated CD4^+^ and CD8^+^ T cells. (F) Percentage of CD38^+^HLA-DR^+^PD-1^+^ triple positive cells in total CD4^+^ and CD8^+^ T subsets. (G) Violin plots showing the expression changes of T cell exhaustion and cytotoxicity-related genes in activated CD4/8^+^ populations across different samples.

### M2 macrophage-polarization and inhibitory pulmonary environment in the severe COVID-19 patient

To elucidate the mechanism causing changes in T cell function and exhaustion post-SARS-CoV-2 infection in the lung, macrophages in PFMC were scrutinized. CCL2 was highly induced in the PFMC macrophages (Figure 2B) and reported to shape macrophage polarization (Sierra-Filardi et al., 2014), so monocytes and macrophages in the COVID-19 patient’s samples were then analyzed. GO term analysis of macrophages confirmed that negative regulators of immune response were selectively enriched in PFMC, while T cell activators were enriched in PBMC and H-PBMC (Figures S3A-S3C). Consistently, anti-inflammatory factors (Stoger et al., 2012), including the M2-macrophage markers, CD163 and CD206 (*MRC1*) (Mantovani et al., 2002), exhibited elevated expression levels on monocytes and macrophages in PFMC, whereas pro-inflammatory marker expression levels were decreased or unaltered (Figures 3A, 3B and S3A). M2 macrophages are key players in T cell exhaustion in the tumor microenvironment and chronic viral infections (Jiang et al., 2015). The potential M2-macrophage polarization event was further validated by elevated expression of M2 polarization regulatory components in PFMC, including CCL2, CSF1, and SPP1 in monocytes as well as APOE, IL10, and CCL18 in macrophages. (Deng et al., 2018; Murray, 2017; Sierra-Filardi et al., 2014; Svensson-Arvelund et al., 2015; Wang et al., 2019; Zhang et al., 2017) (Figures 3C and 3D). APOE and IL-10 are known to induce T cell exhaustion (Deng et al., 2018; Pestka et al., 2004; Yi et al., 2010). Notably, these molecules represented most of the PFMC-specific associations with their receptors upon viral infection (Figures 2A and 2B). Flow cytometry analysis of CD14^+^ monocytes and macrophages in PFMC also demonstrated the presence of CD206^+^ and CD163^+^ populations which were consistent with M2 macrophage phenotype (Figure S3D) (Mantovani et al., 2002). Interestingly, the majority of CD163^+^ cells express PD-1 while CD206^+^ cells express TIM-3, and Poly I:C induced IL-10 expression was primarily found in CD206^+^TIM-3^high^ cells (Figure S3D). Moreover, CCL-2 and IL-10 concentrations were higher in pleural effusion than those in patient plasma, agreeing with the M2 macrophage-driven microenvironment (Figure 3E). Collectively, these results supported the notion that M2 macrophages polarized pulmonary microenvironment potentially contributed to T cell dysfunction in the severe COVID-19 patient (Figure 3F).

**Figure 3.**
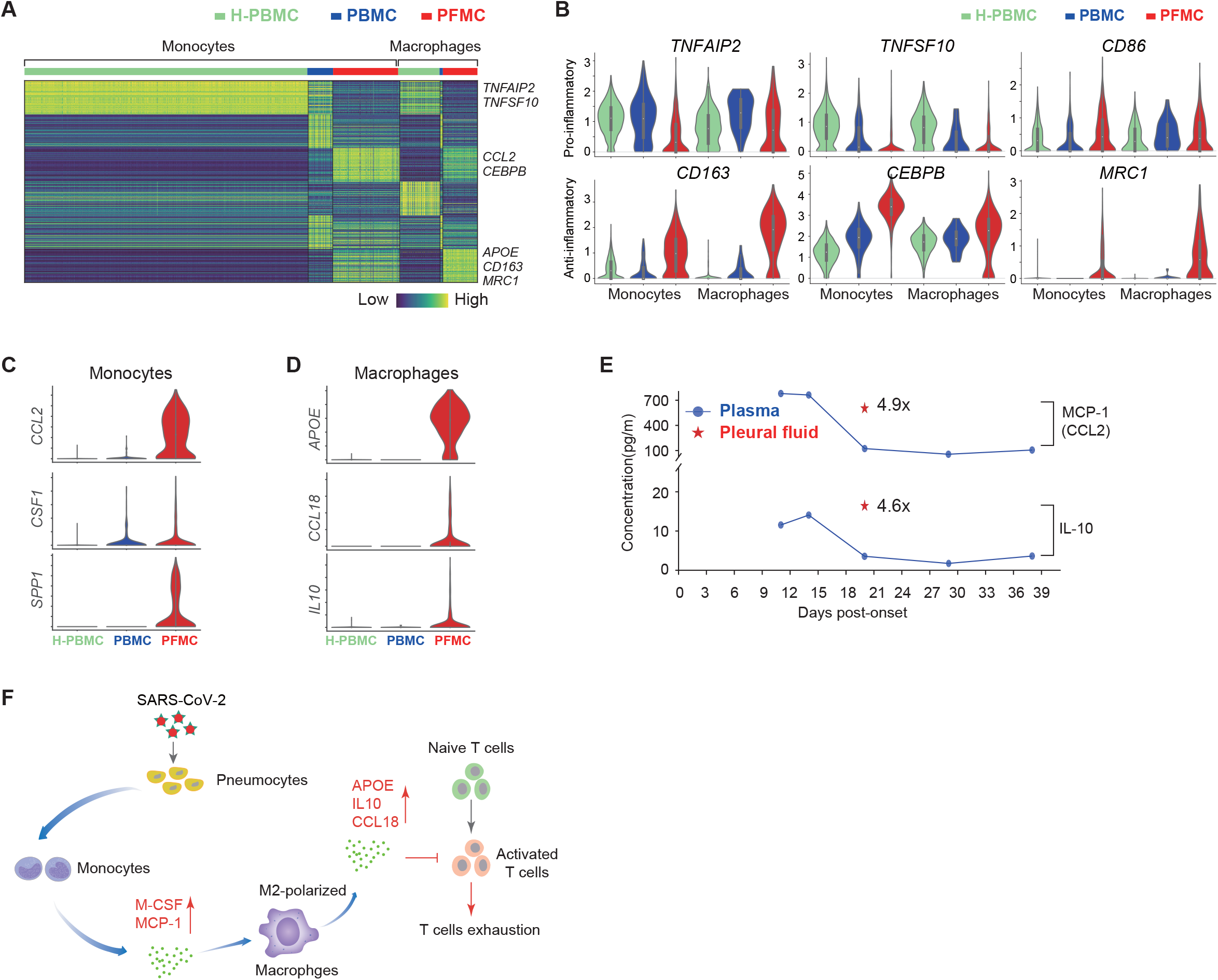
Single-Cell RNA-seq Reveals Unique Macrophage Types Associated with COVID-19 patient. (A) Heatmap showing the differentially expressed genes for macrophages and monocytes between the COVID-19 patient and the healthy donor. (B) Violin plots showing the expression of pro-inflammatory and anti-inflammatory factors between the COVID-19 patient and the healthy donor in the indicated cell types. (C) Violin plots showing the expression of indicated genes in monocytes across different cell types. (D) Violin plots showing the expression of indicated genes in macrophages across different cell types. (E) Concentrations of MCP-1 and IL-10 in the patient’s plasma at the indicated d.p.o. and in the pleural fluid were quantified using BD Cytometric Bead Array (CBA) assay. (F) A model of SARS-CoV-2 pathogenesis. SARS-CoV-2 infects pneumocytes and induces monocyte production of cytokines (MCP-1, M-CSF) that mediate M2 polarization. Activated M2 produce APOE, IL-10 and CCL18 that can inhibit activated T cells and induce T cell exhaustion, leading to the virus immune evasion.

### Characterization of SARS-CoV-2-specific T cell responses in pleural effusion and peripheral blood biopsies of the COVID-19 patient

To characterize virus-specific T cell responses in the patient, PFMCs and PBMCs were stimulated with SARS-CoV-2-specific peptide pools in the presence of brefeldin A. Enhanced number and percentage of viral-specific CD4^+^ and CD8^+^ T cells, determined by IFN-γ expression, were observed in PFMC at 20 *d.p.o*. However, virus-specific T cells were absent in PBMC until later d.p.o. and slowly increased over time (Figures 4A and 4B). Immunocompetent CoV-specific T cells tend to produce multiple cytokines upon peptide stimulation (Zhao et al., 2017). We previously found that in MERS convalescent patients, most of the MERS-CoV-specific T cells co-produced IFN-γ^+^ and TNF^+^ (Zhao et al., 2017). To evaluate the functionality of virus-specific T cells in this patient, co-expression of TNF by IFN-γ^+^ T cells was examined. Most of the virus-specific CD4^+^ and CD8^+^ T cells did not produce TNF in PFMC at 20 d.p.o. and in PBMC at 29 d.p.o. which was consistent with an exhaustion phenotype (Yi et al., 2010) (Figure 4C). The percentage of IFN-γ^+^ TNF^+^ virus-specific T cells increased in PBMC at 38 d.p.o. when the patient’s condition improved (Figure 1A). Of note, most of the IFN-γ^+^ or IFN-γ^+^TNF^+^ cells in PFMC were CD38^+^HLA-DR^+^ which were much higher than those in PBMC (Figure S4A), indicating virus-specific T cells were hyperactivated in the pleural effusion. Finally, the longitudinal elevation of both numbers and functionality of virus-specific T cells were correlated with decreased viral loads in throat swab and sputum specimens (Figure 4D) and disease amelioration (Figure 1A), indicating T cells were required for protection from clinical disease and virus clearance.

**Figure 4.**
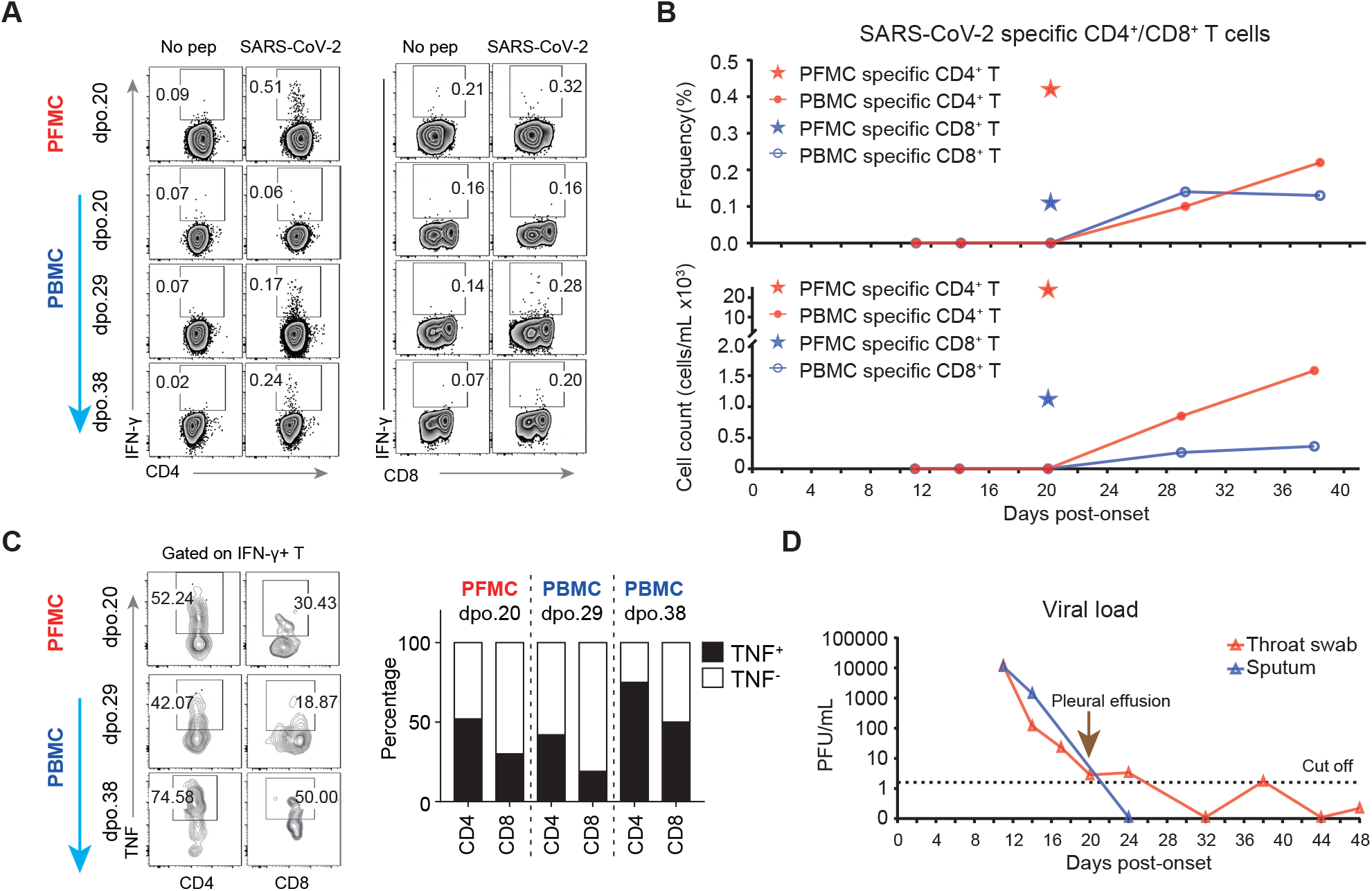
Characterization of virus-specific T cell responses in the pleural fluid and in the peripheral blood. (A) PFMCs and PBMCs obtained at the indicated times post-onset were stimulated with the SARS-CoV-2 peptide pool for 8 hours in the presence of brefeldin A. Virus-specific CD4^+^ and CD8^+^ T cells were identified by intracellular IFN-γ staining. No pep, no peptide control. (B) Frequencies and numbers (cells/mL) of SARS-CoV-2-specific CD4^+^ and CD8^+^ T cells in the patient’s peripheral blood and pleural fluid are shown. (C) Flow cytometry dot plots and a bar graph showing the percentages of virus-specific (IFN-γ+) CD4^+^ and CD8^+^ T cells that co-express TNF. (D) Viral loads in the patient’s sputum and throat swab samples at the indicated times post-onset were shown.

### Corroboration of M2-Polarization in the Sputa of Severe COVID-19 Patients

To validate the macrophage-driven T cell suppression observed in the severe COVID-19 patient’s PFMC, sample size was expanded by including cells isolated from sputum samples from 4 patients, with 2 severe and 2 mild cases (Figure 5A; Table S2). scRNA-seq of these samples revealed 4 major cell types, B cells, epithelia, macrophages, and monocytes (Figures 5B, 5C and S5A). Intriguingly, two samples from severe cases were composed of a larger macrophage population and lower percentages of B cells and epithelia than those in mild patients (Figure 5D). Consistent with PFMC, the anti-inflammatory M2 macrophage markers exhibited higher expression, whereas the pro-inflammatory markers displayed lower mRNA levels in the sputa of the severe COVID-19 patients (Figures 5E, S5B and S5C). Besides, elevated expression of M2-polarization markers further verified the M2 macrophage activation events in the severe cases and highlighted the stage-specific immune response post-SARS-CoV-2 infection (Figure 5F and S5D). Flow cytometry analysis of CD14^+^ leukocytes showed increased M2 macrophages in the severe COVID-19 patient comparing to the mild cases (Figure 5G). In addition, the percentage of M2 cells and M2 polarization related cytokines in sputum were decreased when the severe patient’s condition improved (Figures 5H and 5I), indicating the correlation between M2 cell activation and disease severity.

**Figure 5.**
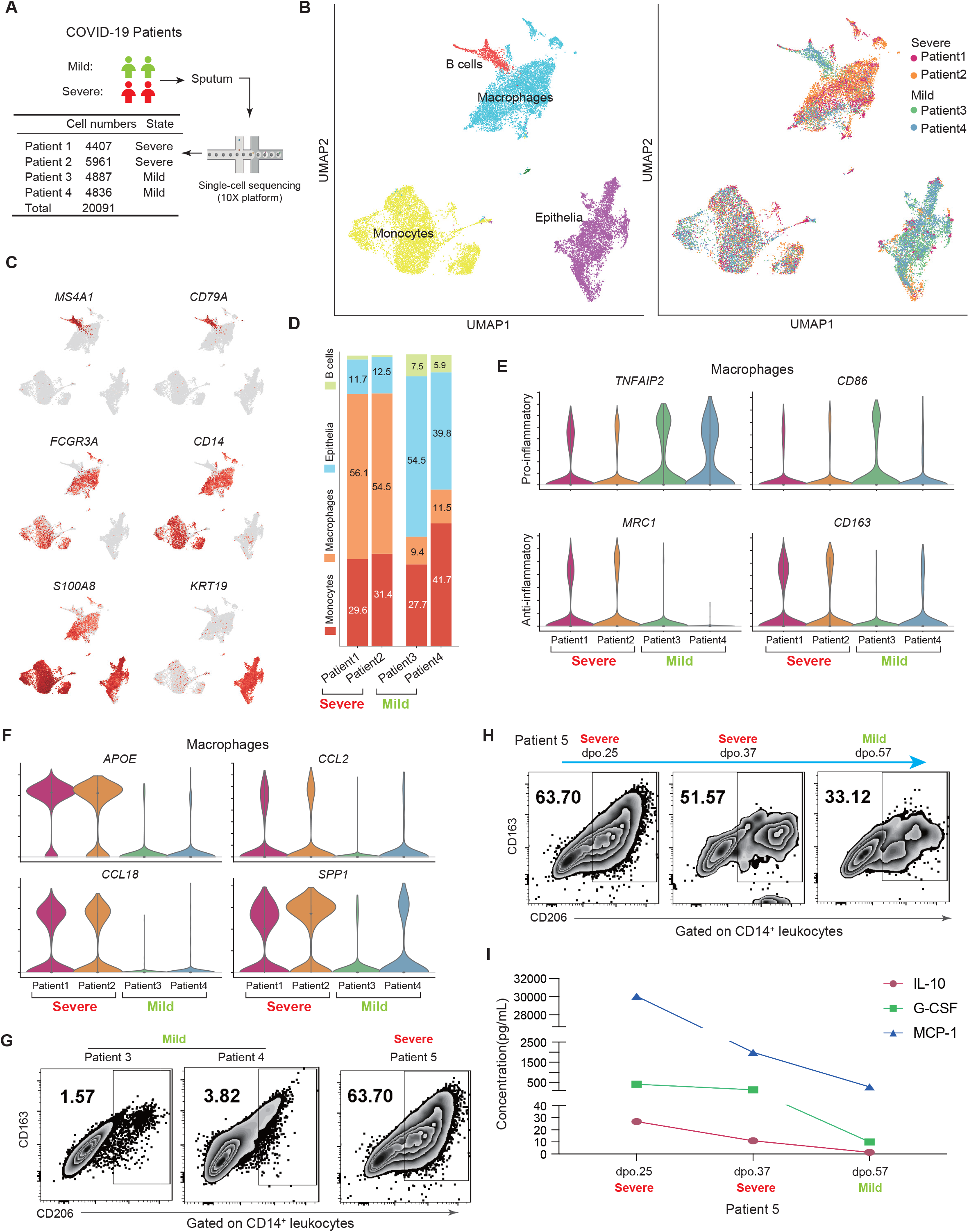
Corroboration of M2-Polarization in the Sputa of Severe COVID-19 Patients. (A) A summary table of the sputum sample information, including sequenced cell numbers and patients’ disease states. Single-cell RNA-seq was performed using the 10X Genomics Chromium system. (B) UMAP plots visualizing a total of 20,091 cells analyzed by single-cell RNA-seq and clustered according to their transcriptomic similarity. Distribution of single cells by cell types (the left panel) and source (the right panel) were displayed. (C) UMAP plots showing the expression of maker genes for all cell types in (B). A higher gene expression level is indicated by a darker shade of red. (D) A 100% stacked column graph displaying the distribution of cell types in each COVID-19 patient. (E) Violin plots demonstrating the expression profiles of pro-inflammatory and anti-inflammatory marker genes in the patients’ macrophages. (F) Violin plots illustrating the expression profiles of M2-polarization genes in the macrophages of each patient. (G) CD14^+^ cells in the severe and mild patient’s sputum were analyzed for M2 macrophage markers CD206 and CD163. (H) Sputum obtained at the indicated times post-onset were analyzed for M2 macrophage markers CD206 and CD163. (I) M2 polarization related cytokines (IL-10, G-CSF and MCP-1) in sputum were analyzed at the indicated times post-onset.

## DISCUSSION

SARS-CoV-2 is the causative pathogen of COVID-19 pneumonia (Zhou et al., 2020). The elderly and individuals with comorbidities are at higher risk for severe disease (Guan et al., 2020; Li et al., 2020b). No drug or vaccine has yet been approved for human use due to the lack of understanding of disease pathogenesis and immune responses in human.

Our study provides a framework to systematically investigate immune profile in COVID-19 patient-derived PFMC and sputum biopsies. By integrating advanced single-cell technology and immunological approaches, M2-macrophage polarization-driven T cell exhaustion upon viral infection in the lung was mechanistically revealed suggesting T cell suppression and dysregulation in infection site of severe COVID-19 cases. SARS-CoV-2-specific T cells were also detected in PFMC earlier than in PBMC of the patient, demonstrating the advantage of utilizing PFMC biopsies. The sensitivity and reliability of PFMC over PBMC had been underlined throughout this study. In addition to the earlier emergence of the SARS-CoV-2-specific T cell population, PFMC also encompassed a higher number of hyperactivated T cells. A group of CD14^+^ M2 macrophage-like cells highly expressing M2 polarization markers, and as well as T cells expressing exhaustion markers were also exclusively found in PFMC. Comparing to the subtle hints from PBMC, PFMC served as a robust tool and provided consistent mechanistic evidence of immune dysregulation.

As excess fluids in the pleural cavity surrounding the lungs, pleural effusion had been found to associate with severe pulmonary infections, including MERS (Das et al., 2015) and COVID-19 (Shi et al., 2020). Pulmonary pathological microenvironment had been reported to increase the permeability of the capillaries in the lung, leading to exudate pleural effusion (Porcel and Light, 2006, 2008). Thus, PFMC was reasoned to share closer immune properties in the SARS-CoV-2-infected lungs in human and a better indicator of the pulmonary inflammatory microenvironment. Further, post-mortem examination of a severe COVID-19 case illustrated thickened pleura with extensive adhesion to the lung tissue (Liu et al., 2020b) suggesting the involvement of pleura and pleural fluids in disease pathogenesis. These evidences supported our conclusion and highlighted the translational value of this study.

The cutting-edge single-cell analysis of the PFMC biopsy facilitated the profiling of the heterogeneous immune response in the patient. Through ligand-receptor interaction analysis, enhanced T cell activation and exhaustion pathways were accompanied by the elevation of their mRNA expression levels. Furthermore, augmented M2 polarization markers in the monocytes and macrophages mechanistically linked T cell exhaustion with M2 macrophage activation, reflecting virus-induced T cell suppression, which is consistent with our previous finding that alveolar macrophages play inhibitory roles on virus-specific T cells in SARS-CoV infected mice (Zhao et al., 2009). Although prevailing perspectives had associated exuberant inflammatory response with severe CoV cases (Mahallawi et al., 2018; Wong et al., 2004), COVID-19 exhibited distinct pathological phenotypes indicating it has distinct immune properties after infection. In contrast to SARS, the COVID-19 autopsy displayed less fibrosis, suggesting a dampened release of pro-inflammatory factors (Biswas et al., 2011). Huang *et al*. also reported the increased secretion of immunosuppressive cytokines, IL-4 and IL-10, demonstrating a COVID-19-specific cytokine profile (Huang et al., 2020). In a mouse model of SARS-CoV infection, STAT-I was also found to induce M2 polarization, causing pulmonary damage (Page et al., 2012). These pathological and immunological analyses supported that immune dysregulation and dysfunction contributed to COVID-19-specific pathogenesis, echoing our observations in the severe COVID-19 cases.

Our findings unveiled that the immunoinhibitory pulmonary environment driven by M2 macrophage polarization induced T cell suppression in severe COVID-19 patients, suggesting anti-M2 treatment as a potential therapeutic strategy. Some approved anti-M2 strategy has already achieved good clinical outcomes in anti-tumor therapy (Tariq et al., 2017). Agents such as targeting of IL-10, CCL2, and CSF signaling axis have demonstrated efficacy in preclinical breast cancer (Germano et al., 2013; Tariq et al., 2017; Zollo et al., 2012). Alternatively, enhancing M1 with anti-CD40 or CD25 monoclonal antibody (mAb) to suppress M2 polarization has also been reported (Buhtoiarov et al., 2005; Jacobs et al., 2010; Tariq et al., 2017). Besides, severe COVID-19 patients in our hospital were regularly treated with Thymosin (Zadaxin), a T cell stimulator which has been shown to stimulate T cell development, differentiation and proliferation (Costantini et al., 2019; Garaci et al., 1995; Jiang et al., 2011). After Zadaxin treatment, increased NK and CD8 T cell numbers and partially reversed T cell exhaustion were observed, indicating Zadaxin could potentially restore T cell function (Figures S1C and Fig 4C).

Lymphopenia had been considered as a signature of severe COVID-19 infection (Bermejo-Martin et al., 2020). Meanwhile, impaired lymphatic tissue in spleen and lymph node shrinkage was observed in a COVID-19 autopsies (Liu et al., 2020b), indicating massive damage of immune system. Integrating our findings, to maintain the proliferation and functionality of T cells before the massive immune suppression would prevent the deterioration of disease. Conversely, the progression of T cell suppression could lead to excessive T cell exhaustion, and consequently, result in delayed viral clearance.

In conclusion, our study utilized the pleural effusion and sputum biopsies derived from severe COVID-19 cases to dissect the heterogeneity of immune response upon SARS-CoV-2 infection in human. Specifically, strong mechanistic evidence of M2 macrophage-driven T cell exhaustion was elucidated. Further analysis of the correlation between T cell suppression and disease development demonstrated that the value of pleural effusion and sputa in translational research and providing mechanistic implications for disease diagnostics and treatment.

## Data Availability

Data are available upon reasonable request

---

**Figure S1.**
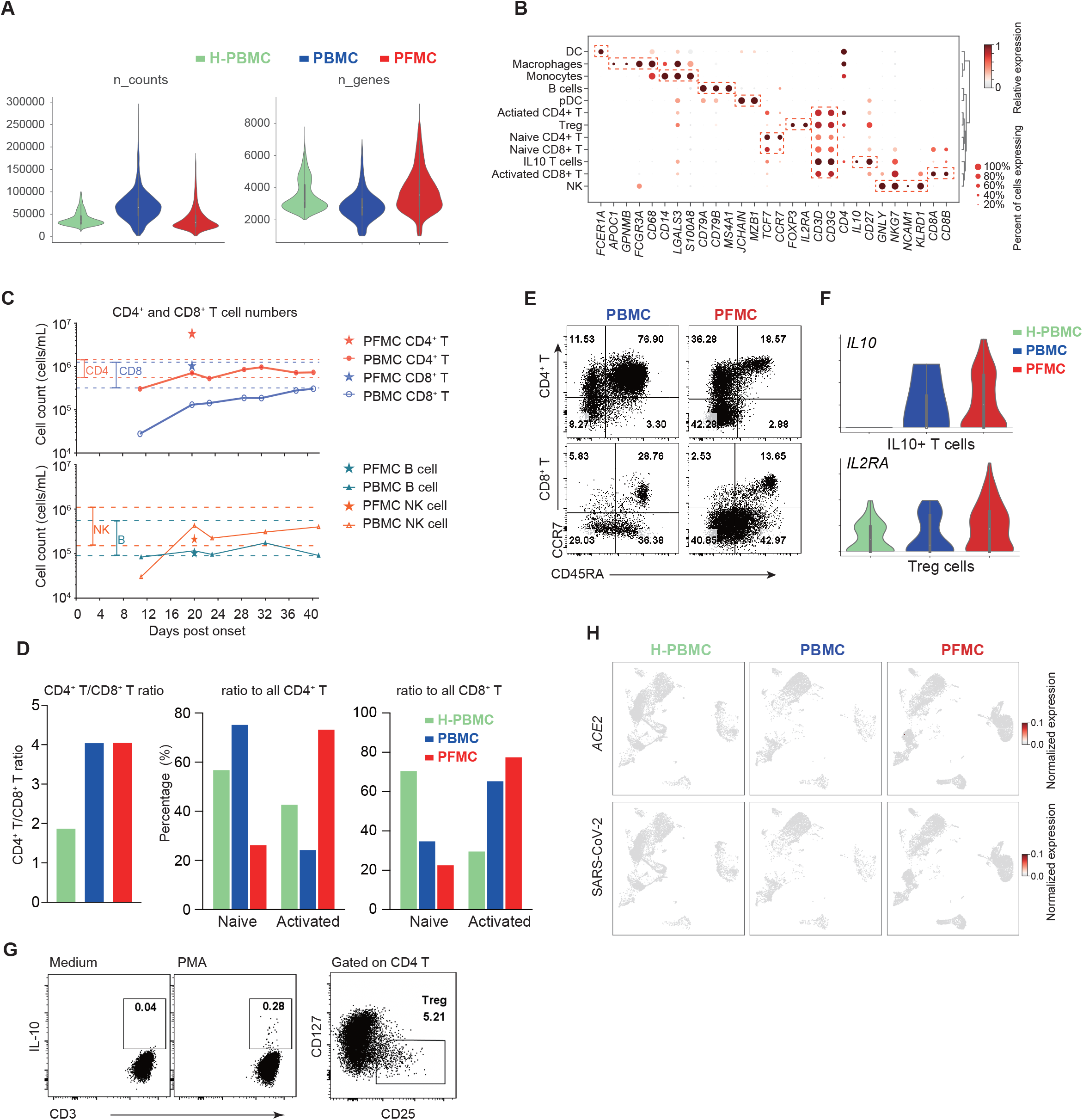
Identification of Cell Types for COVID-19 Patient. (A) Violin plots showing the raw counts number and the numbers of detected genes by scRNA-seq for each cell. (B) Dot plots showing the expression level and the percent of cells the gene is expressed in, for the indicated marker genes. (C) Absolute CD4/8^+^ T cell (upper) and NK/B cell (lower) counts (cells/mL) in the patients’ pleural fluid and peripheral blood collected at the indicated times post-onset are shown. The red and blue dashed lines represent the normal range of CD4^+^ and CD8^+^ T cell counts respectively. (D) Bar graphs showing the indicated ratios across samples. (E) Flow plots showing CCR7 and CD45RA expression by CD4^+^ and CD8^+^ T cells from the patient’s PBMC and PFMC. (F) Violin plots showing the expression of IL2RA in Treg cells (upper) and IL10 in IL10+ T cells (lower) defined from Figure 1C. (G) (left) PFMCs were stimulated with PMA and ionomycin for 4 hours in the presence of brefeldin A. IL-10 expression by a small population of T cells were detected by intracellular cytokine staining; (right) A flow pot showing CD4^+^CD25^+^CD127^low^ regulatory T cells present in the PFMC. (H) UMAP plots showing the expression of *ACE2* and SARS-CoV-2. Gene expression levels are indicated by shades of red.

**Figure S2.**
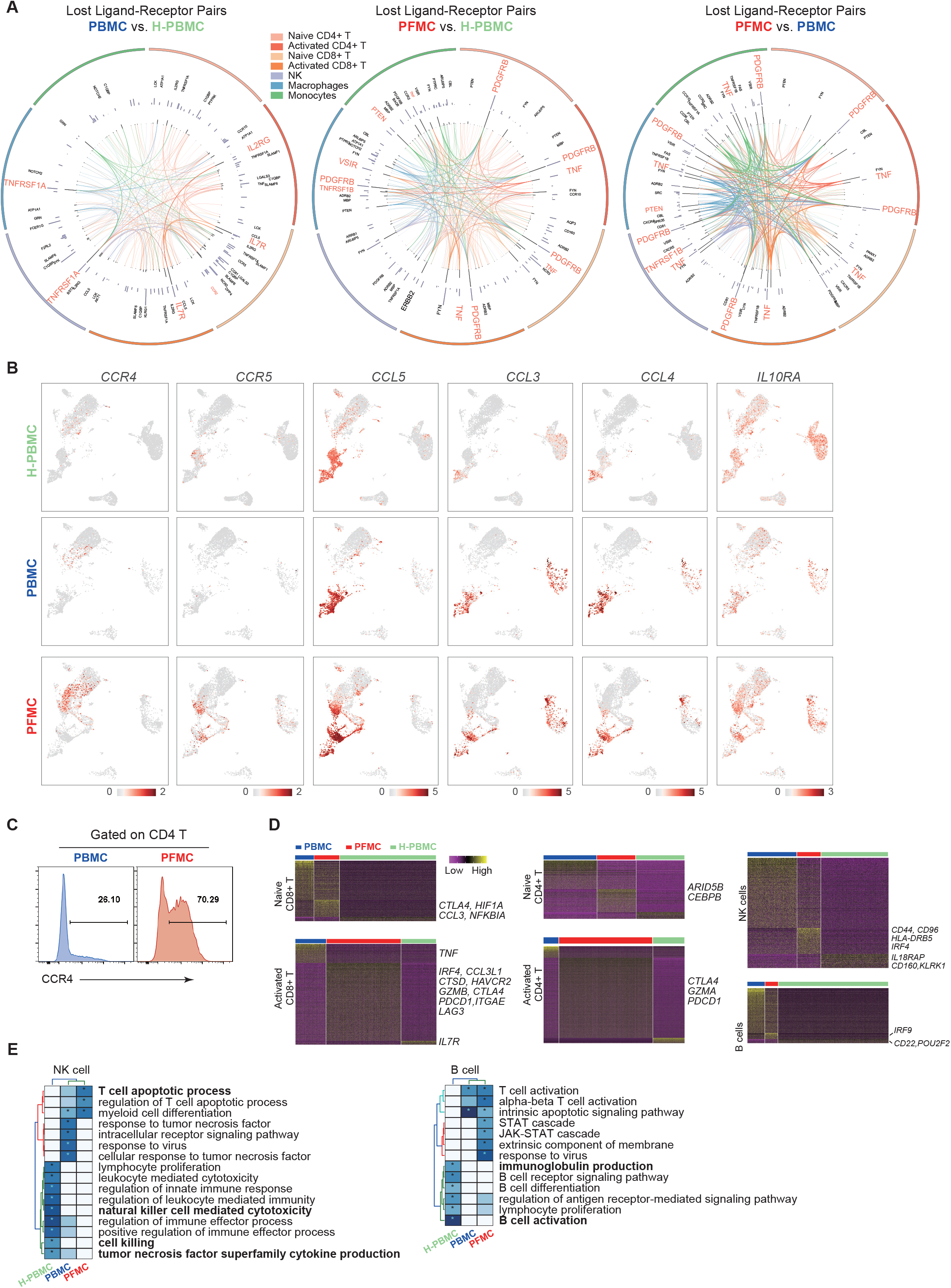
Virus-induced responses in COVID-19 patient. (A) Circos plots showing the ligand-receptor interactions across different cell types. The outermost ring represents different cell types according to the color bar. The innermost color lines represent ligand-receptor contacts in indicated cell types, the line thickness represents the contact strength. The adjacent black ring shows ligand-receptor interaction strength changes in between indicated samples. The next black ring represents the expression changes of indicated genes in the indicated cell types. (B) UMAP plots showing the expression of indicated genes, Gene expression levels are indicated by shades of red. (C) Flow histograms showing CCR4 expression by CD4^+^ T cells. (D) Heatmaps showing the differentially expressed genes in indicated cell types across different cell types. Selected genes are indicated on the right. (E) Gene ontology (GO) analysis for the differential expressed genes from panel D.

**Figure S3.**
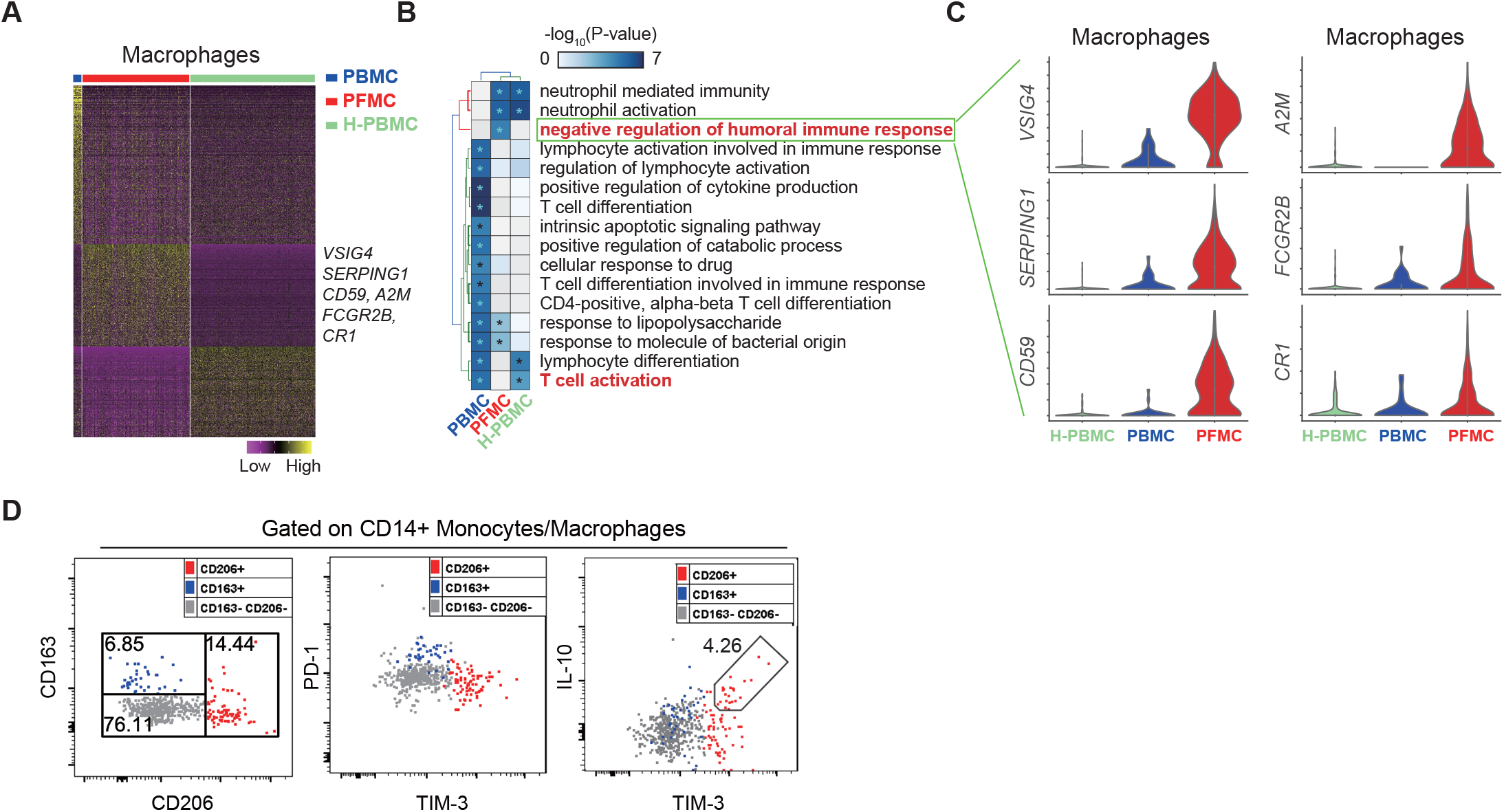
Unique macrophages in COVID-19 patient. (A) Heatmap showing the differentially expressed genes in macrophages from different cell origins. Selected genes are indicated on the right. (B) Gene ontology analysis for PFMC specifically high expression genes from panel A. (C) Violin plots showing the differential expression of humoral immune response negative regulation-related genes in macrophages between difference samples. (D) CD14^+^ cells in the patient’s pleural fluid were analyzed for M2 macrophage markers CD206 and CD163 as well as inhibitory markers PD-1 and TIM-3. IL-10 was detectable in TIM-3^high^ CD14^+^ cells when PFMCs were stimulated with Poly I:C for 4 hours in the presence of brefeldin A.

**Figure S4.**
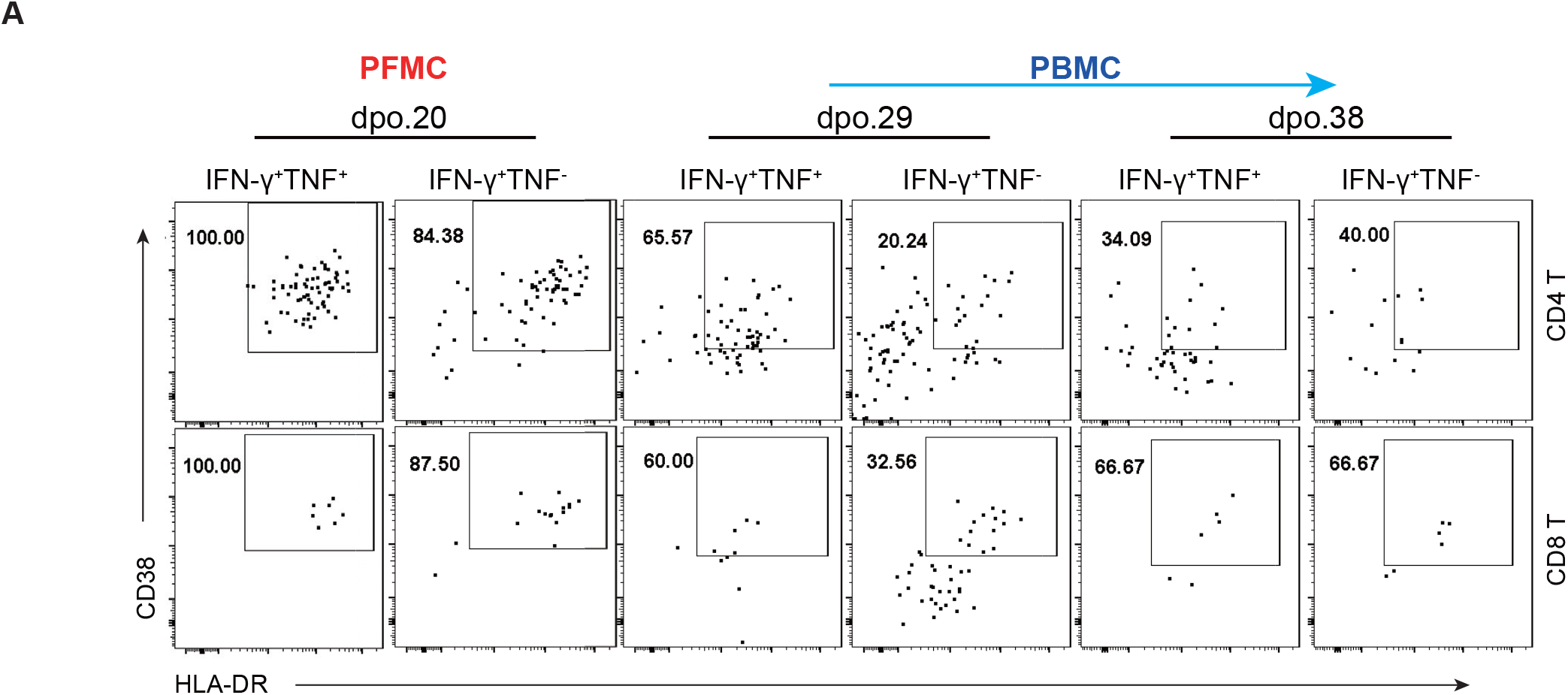
Anti-viral T Cell Responses. (A) Flow plots showing CD38 and HLA-DR expression by IFN-γ^+^TNF^+^ and IFN-γ^+^TNF^-^ virus-specific CD4^+^ and CD8^+^ T cells from the patient’s pleural fluid and peripheral blood.

**Figure S5.**
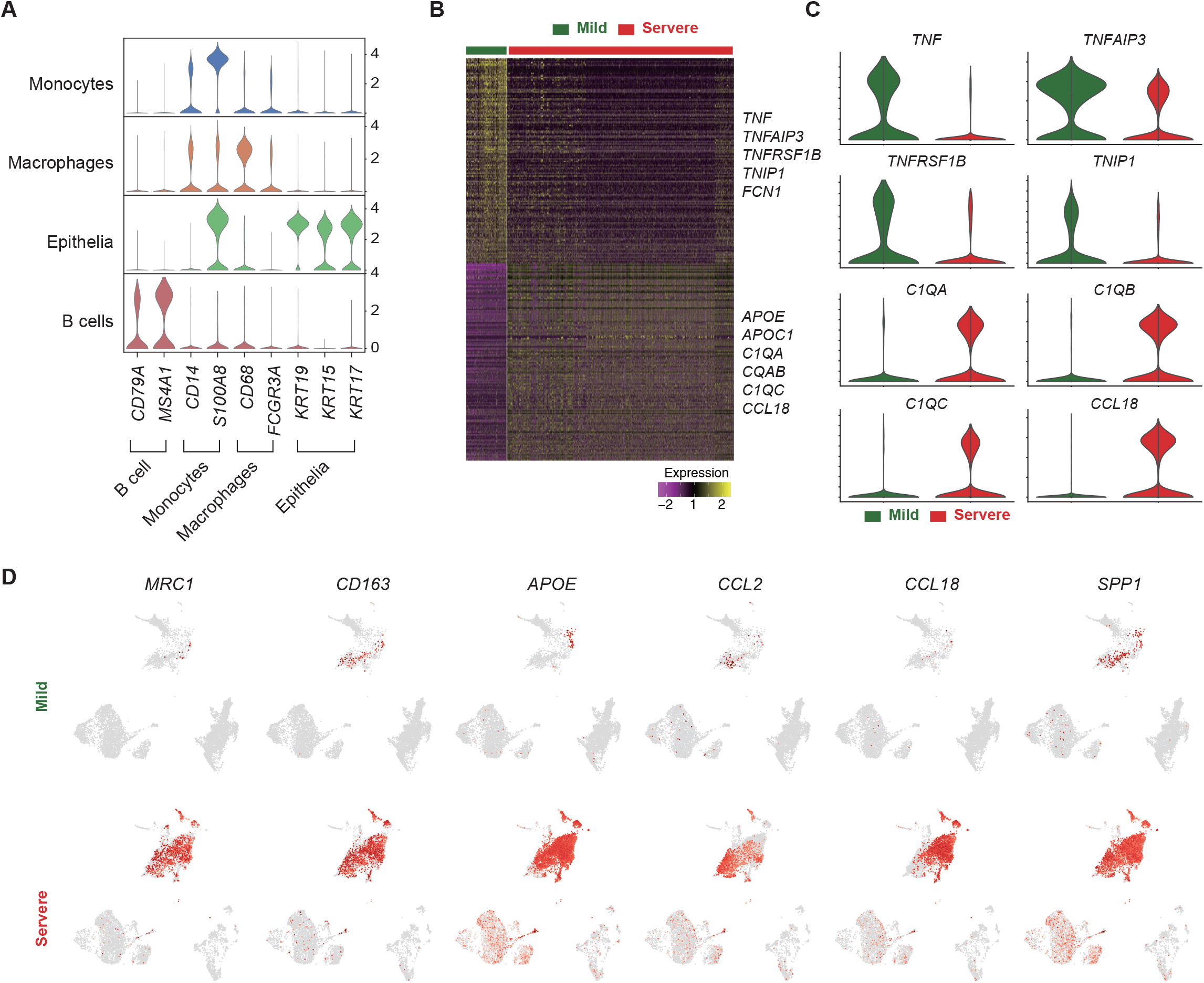
Single-cell Transcriptomic Profiles of Sputum Samples From 4 COVID-19 Patients. (A) Violin plots portraying the expression of classic markers for each identified cell types, validating the cell type annotation in Figure 5B. (B) A heatmap showing the differentially expressed genes in the macrophages of the mild (green) and severe (red) COVID-19 cases. Yellow and purple colors represent high and low expression levels, respectively. Selected genes are highlighted on the right. (C) Violin plots displaying the expression of pro-inflammatory (i.e., TNF, TNFAIP3, TNFRSF1B, and TNIP1) and anti-inflammatory (i.e., C1QA, C1QB, C1QC, and CCL18) markers in the macrophages of mild (green) and severe (red) COVID-19 patients. (D) UMAP plots showing the expression of M2-polarization genes. A higher gene expression level is indicated by a darker shade of red.

## EXPERIMENTAL PROCEDURES

### Patient information

A 70s old man was laboratory-confirmed positive for novel coronavirus (SARS-CoV-2) infection by real-time PCR. After a 9-day treatment in an isolation ward, the patient became critically ill with respiratory failure and multiple organ dysfunctions, and was transferred to the intensive care unit (ICU) of the First Affiliated Hospital of Guangzhou Medical University on Feb 3, 2020. Notably, pleural effusion was observed on Feb 13 and a thoracentesis was performed under ultrasound guidance and a draining tube was placed on Feb 14. The respiratory support for this patient was successfully switched from invasive mechanical ventilation to non-invasive intermittent positive pressure ventilation on March 1. The patient is still in ICU and gradually recovering now. Additional 3 severe patients and 2 mild patients were also included in this study.

### Study approval

This study was approved by the Ethics Committee of the First Affiliated Hospital of Guangzhou Medical University with written consent form acquired from the patient. Patient samples were obtained at the indicated times and clinical information was retrieved from the clinical record.

### Sample collection and preparation

Throat swabs, sputum, peripheral blood and pleural fluid samples were collected by professional nurses in the hospital and were processed in a biosafety level 2 plus laboratory with biosafety level 3 personal protection equipment. For RNA detection, 3 mL DMEM medium containing 2% FBS was added to swabs and sputum tubes, and following 2500 rpm, 15-30 sec vortex and 15-30 min standing, the supernatant was collected and added to lysis buffer for RNA extraction. Supernatant from pleural fluid was collected following 400 g, 5 min centrifugation for RNA extraction or cytokine detection, and cells in the pleural fluid (PFMC) were freshly used for 10x single-cell RNA-seq or cryopreserved in liquid nitrogen. Sputa were washed once with equal volume of PBS, supernatant was collected for cytokine analysis. Sputa were then treated with equal volume of 0.2% DTT in PBS and cells were collected by centrifugation. Plasma was collected from peripheral blood and PBMCs were isolated using Leucosep tubes (Greiner) and Ficoll-Paque PLUS (GE Healthcare) according to the manufacturer’s instructions. Plasma was stored at −80°C for cytokine detection and PBMCs were freshly used for 10x single cell RNA seq or stored in liquid nitrogen.

### Viral load quantification

Nucleic acid of respiratory samples was extracted using a Viral RNA extraction kit from Zybio Inc (Chongqing). An in-house real-time PCR kit targeting the SARS-CoV-2 orf1ab gene region was provided by Zybio Inc. Viral loads in respiratory specimens were measured by qRT-PCR and standardized as PFU/ml relative to a positive control (authentic SARS-CoV-2) with known infectious viral titer.

### Cytokine quantification

Cytokine levels in plasma, sputum and pleural fluid were quantified using BD Cytometric Bead Array (CBA) according to the manufacture’s protocol with slight modification. Briefly, 50 μL sample was incubated with 50 μL mixed capture beads and 50 μL detection reagent at RT for 3 hours in the dark. Beads were then washed once with 1 mL wash buffer by centrifugation at 300 g for 5 min. Beads were ultimately suspended in the BD Cytofix fixation buffer instead of Assay Diluent. A single set of diluted standards was used to generate a standard curve for each analyte. Flow cytometry data were acquired on a BD FACSVerse flow cytometer and analyzed using FCAP v3.0 software.

### Absolute cell count

To determine absolute numbers of leukocyte subpopulations in blood, BD Trucount tubes (BD, Catalog No. 340334) were used. Briefly, a 20 μL fluorescence-conjugated antibody cocktail was added to the Trucount tube, followed by 50 μL fresh whole blood, mixed well and incubated for 15 min at room temperature (RT) in the dark. Then 450 μL BD FACS lysing solution (BD, Catalog No. 349202) was added and incubated for 15 min at RT to lyse red blood cells. Samples were analyzed on a FACSVerse flow cytometer.

### SARS-CoV-2 peptide library

To generate a SARS-CoV-2 peptide library, the sequence from a reference strain, BetaCoV/Wuhan/IVDC-HB-01/2019 (Accession No. EPI_ISL_402119) isolated from a patient during the early SARS-CoV-2 pandemic in Wuhan, China was used. A total of two hundred and eighty 20-mer peptides overlapping by 10 amino acids, covering the four SARS-CoV-2 structural proteins, including the spike (S) glycoprotein, the nucleocapsid (N) protein, and the transmembrane (M) and envelope (E) proteins, were synthesized and used for stimulation of PBMCs and PFMCs *in vitro*. Virus-specific T cell responses were subsequently determined using intracellular cytokine staining assays for interferon-gamma (IFN-γ) and tumor necrosis factor (TNF) expression.

### Flow cytometry

For surface staining, 10^5^ to 10^6^ cells were blocked with Fc receptor blocking solution (Biolegend), stained with the indicated antibodies at RT for 15 min, labeled with LIVE/DEAD staining dye (Thermo Fisher), and then fixed with Cytofix Solution (BD Biosciences). For intracellular cytokine staining, 10^5^ to 10^6^ cells per well were cultured in 96-well round-bottom plates at 37°C for indicated times in the presence of brefeldin A (BD Biosciences) and stimulators as follows: for virus-specific T cell stimulation, SARS-CoV-2 peptide pool (0.15~0.3 μM for each individual peptide, GenScript) was used; for bulk T cell stimulation, 50 ng/mL PMA (SIGMA) + 1μg/mL ionomycin (abcam) was used; for macrophage stimulation, 1μg/mL Poly (I:C) (Invivogen) was used to mimic viral RNA. Cells were then labeled for cell surface markers, fixed/ permeabilized with Cytofix/Cytoperm Solution (BD Biosciences), and labeled with anti-intracellular cytokine/protein antibodies. Flow cytometry data were acquired on a BD FACSVerse or a BD Aria III flow cytometer and analyzed using FlowJo software (Tree Star Inc.).

The following anti-human monoclonal antibodies were used:

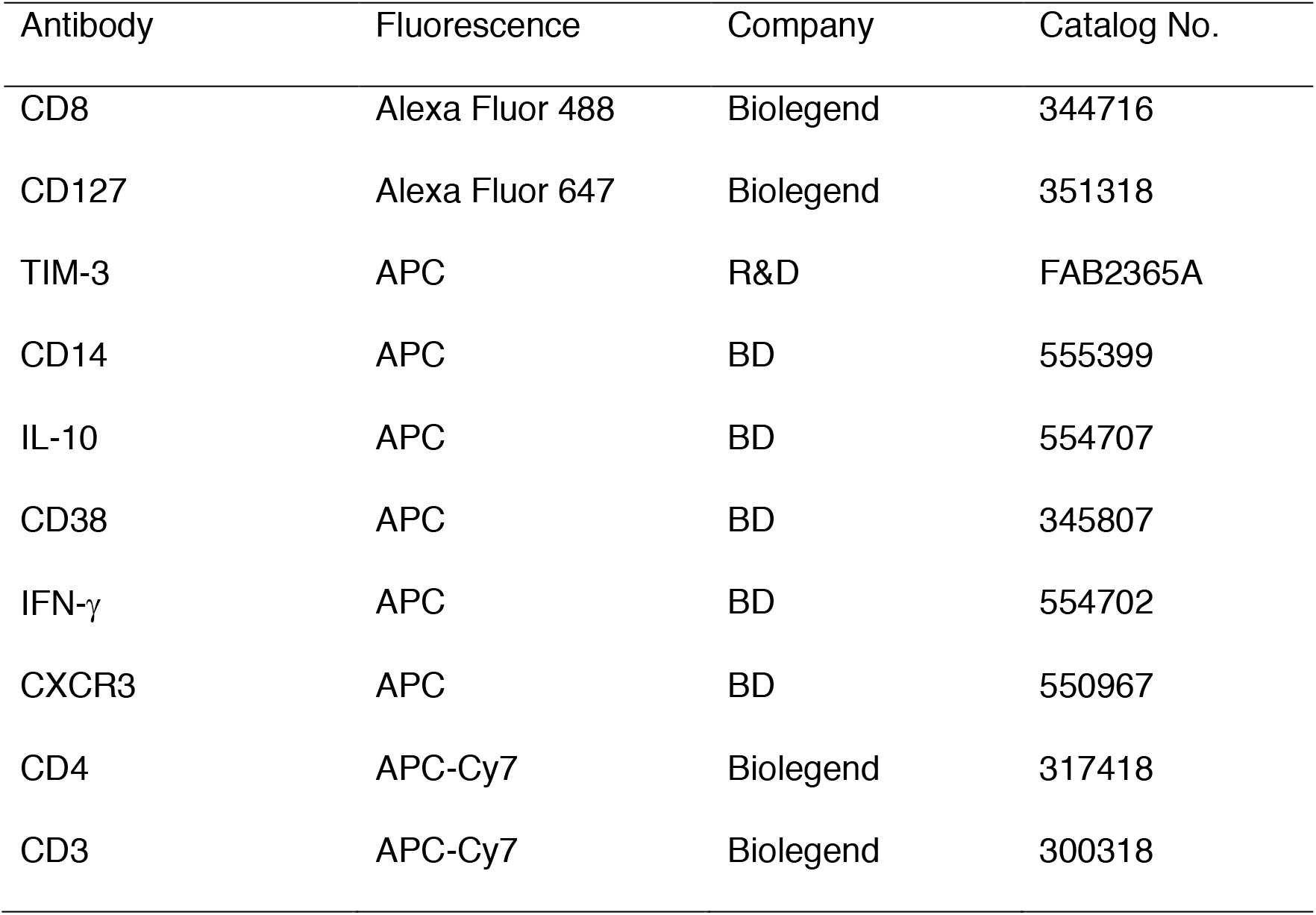
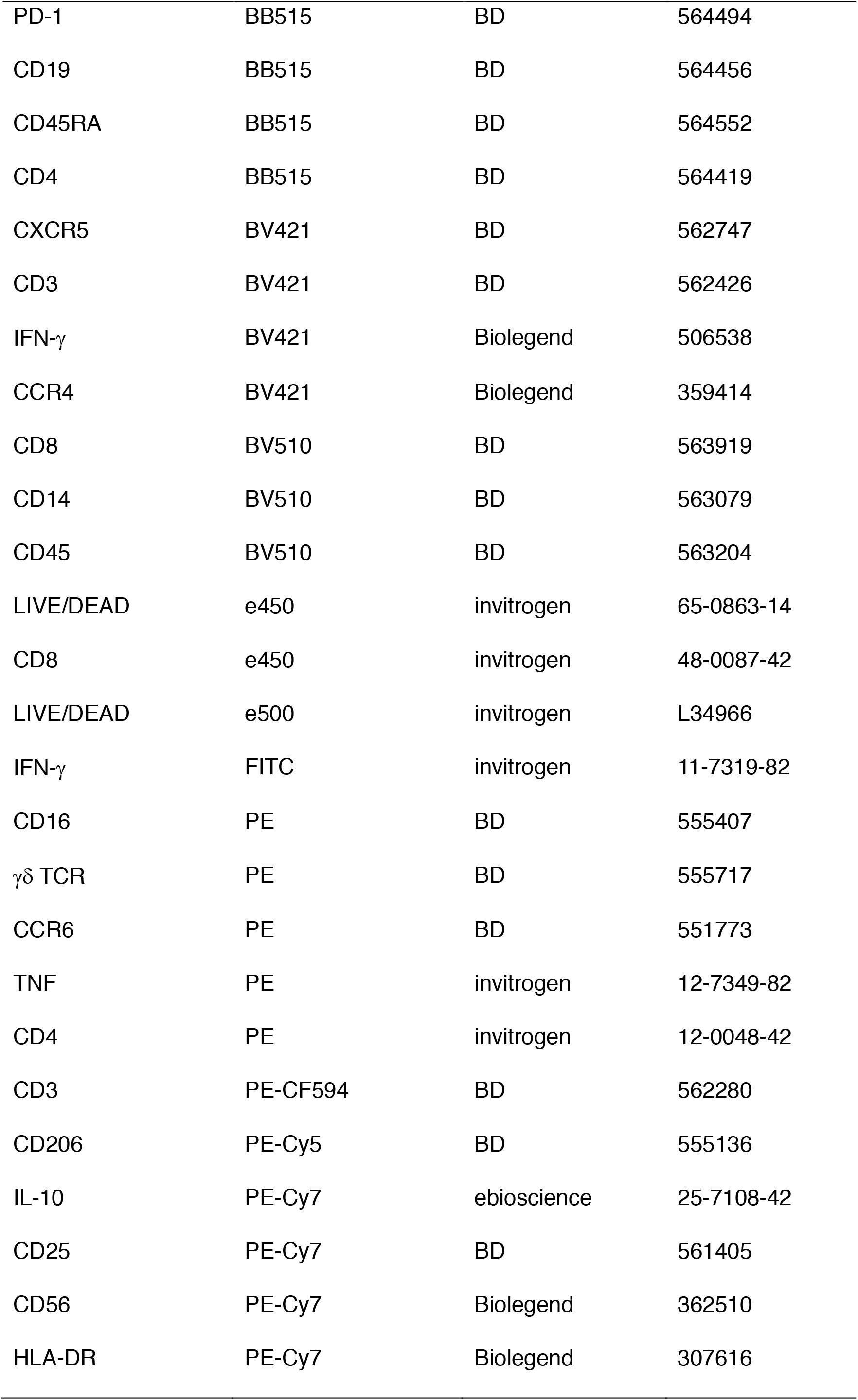
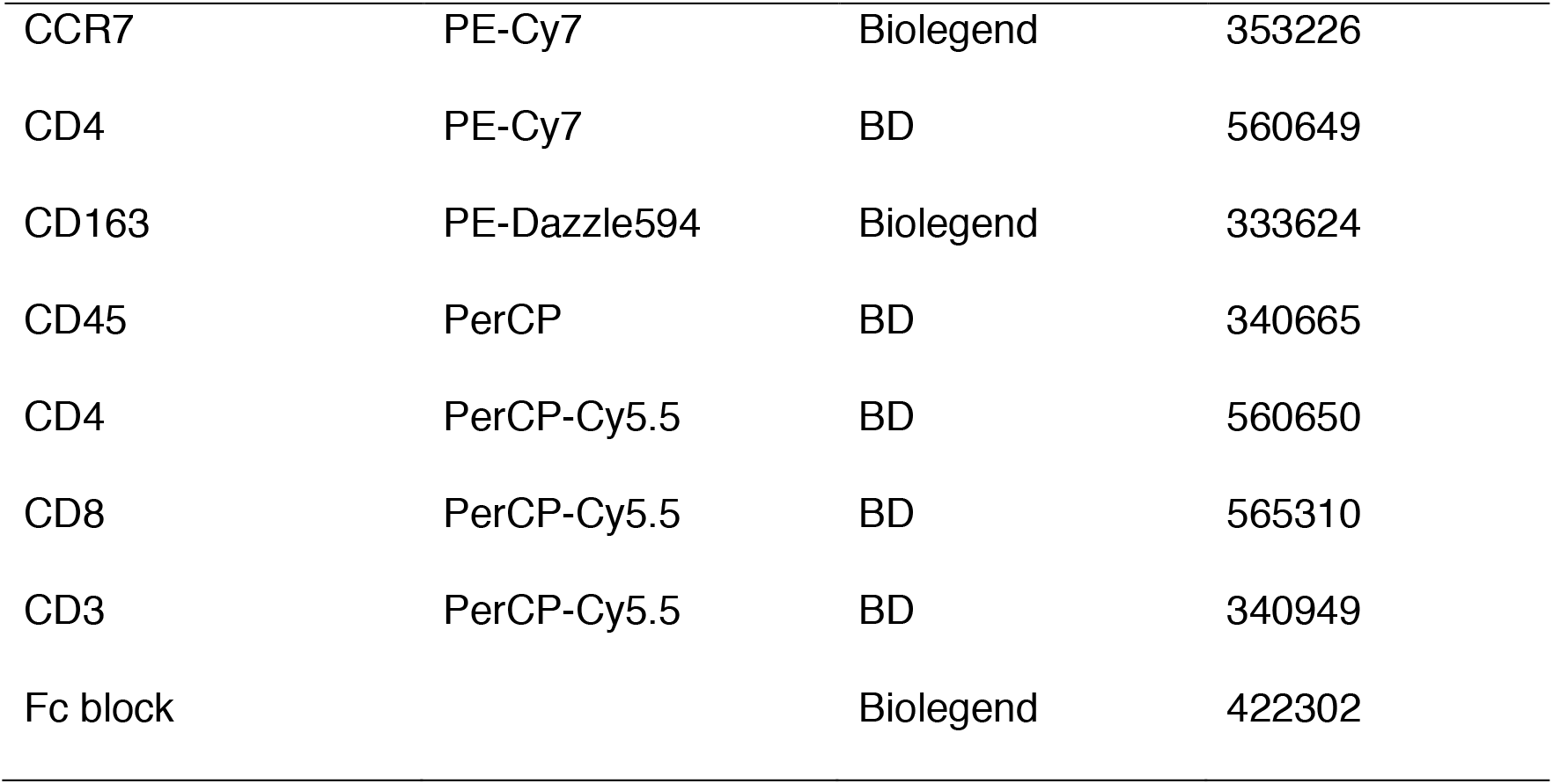

| Antibody | Fluorescence | Company | Catalog No. |
| --- | --- | --- | --- |
| CD8 | Alexa Fluor 488 | Biolegend | 344716 |
| CD127 | Alexa Fluor 647 | Biolegend | 351318 |
| TIM-3 | APC | R&D | FAB2365A |
| CD14 | APC | BD | 555399 |
| IL-10 | APC | BD | 554707 |
| CD38 | APC | BD | 345807 |
| IFN-γ | APC | BD | 554702 |
| CXCR3 | APC | BD | 550967 |
| CD4 | APC-Cy7 | Biolegend | 317418 |
| CD3 | APC-Cy7 | Biolegend | 300318 |
| PD-1 | BB515 | BD | 564494 |
| CD19 | BB515 | BD | 564456 |
| CD45RA | BB515 | BD | 564552 |
| CD4 | BB515 | BD | 564419 |
| CXCR5 | BV421 | BD | 562747 |
| CD3 | BV421 | BD | 562426 |
| IFN-γ | BV421 | Biolegend | 506538 |
| CCR4 | BV421 | Biolegend | 359414 |
| CD8 | BV510 | BD | 563919 |
| CD14 | BV510 | BD | 563079 |
| CD45 | BV510 | BD | 563204 |
| LIVE/DEAD | e450 | invitrogen | 65-0863-14 |
| CD8 | e450 | invitrogen | 48-0087-42 |
| LIVE/DEAD | e500 | invitrogen | L34966 |
| IFN-γ | FITC | invitrogen | 11-7319-82 |
| CD16 | PE | BD | 555407 |
| $\gamma\delta$ TCR | PE | BD | 555717 |
| CCR6 | PE | BD | 551773 |
| TNF | PE | invitrogen | 12-7349-82 |
| CD4 | PE | invitrogen | 12-0048-42 |
| CD3 | PE-CF594 | BD | 562280 |
| CD206 | PE-Cy5 | BD | 555136 |
| IL-10 | PE-Cy7 | ebioscience | 25-7108-42 |
| CD25 | PE-Cy7 | BD | 561405 |
| CD56 | PE-Cy7 | Biolegend | 362510 |
| HLA-DR | PE-Cy7 | Biolegend | 307616 |
| CCR7 | PE-Cy7 | Biolegend | 353226 |
| CD4 | PE-Cy7 | BD | 560649 |
| CD163 | PE-Dazzle594 | Biolegend | 333624 |
| CD45 | PerCP | BD | 340665 |
| CD4 | PerCP-Cy5.5 | BD | 560650 |
| CD8 | PerCP-Cy5.5 | BD | 565310 |
| CD3 | PerCP-Cy5.5 | BD | 340949 |
| Fc block |  | Biolegend | 422302 |

The following Cytometric Bead Array (CBA) products were used to detect cytokines:

| Cytokine | Company | Catalog No. |
| --- | --- | --- |
| Human Th1/Th2/Th17 kit (IL-10) | BD | 560484 |
| Human MCP-1 Flex Set | BD | 558287 |

### Single-Cell RNA library preparation and sequencing

Cell suspensions were loaded onto a chromium single-cell chip to generate single-cell gel bead-in-emulsions (GEMs) aiming for 2,000-8,000 single cells per reaction. The single-cell 3’-library was constructed using Chromium Single Cell Reagent Kits v3 (10X GENOMICS) following the manufacturer’s user guide. Following cell lysis, first-strand cDNA synthesis and amplification were carried out according to the instructions. Amplified cDNA was purified using SPRIselect beads (Beckman Coulter) and sheared to 250-400 bp. cDNA quality control was performed using Qubit 3.0 Fluorometer and Agilent Bioanalyzer 2100. The linear DNA libraries were converted to a single-stranded circular(ssCir) DNA library by MGI Easy Universal Library Conversion Kit (App-A, MGI) and sequenced on BGISEQ-500 with high-throughput sequencing set (App-A, MGI) with the following read lengths: 28-bp read 1 (containing the 18-bp cell barcode and 10-bp randomer), 100-bp read 2 and 8-bp barcodes.

### Single-cell RNA-seq data analysis

scRNA-seq data was processed using the scTE (https://github.com/jphe/scTE) 10x pipeline, Briefly, reads were aligned to the human genome (hg38) using STARsolo(Dobin et al., 2013) with the setting ‘--outSAMattributes NH HI AS nM CR CY UR UY --readFilesCommand zcat --outFilterMultimapNmax 100-- winAnchorMultimapNmax 100 --outMultimapperOrder Random --runRNGseed 777 --outSAMmultNmax 1’. The default scTE parameters for 10x were used to get the molecule count matrix. The count matrix was lightly filtered to exclude cell barcodes with low numbers of counts: Cells with less than 1000 UMIs, less than 500 genes detected or more than 20% fraction of mitochondrial counts were removed. For comparison between patient PFMC, PBMC and control healthy PBMC, the batch effect was corrected by Seurat (V3)(Stuart et al., 2019). The genes with fold change >1.5 and adjusted P-value <0.01 (Wilcoxon test) were considered to be differentially expressed. The Gene Ontology (GO) analysis was performed by clusterProfiler(Yu et al., 2012). Other analysis was performed by SCANPY(Wolf et al., 2018).

### Ligand-receptor interaction analysis

The ligand-receptor interacting patterns annotation were downloaded from iMEX consortium (http://www.imexconsortium.org/) (Orchard et al., 2012) and CellPhoneDB (https://www.cellphonedb) (Vento-Tormo et al., 2018), we excluded from our analysis with only secreted, cytokines, growth factors, hormones, extracellular matrix, membrane, receptors and transporters according to uniport classification. The cell-cell communication analysis followed the previous pipeline (Vento-Tormo et al., 2018). We randomly permuted the cluster labels of all cells 1,000 times and determined the mean of the average receptor expression level of a cluster and the average ligand expression level of the interacting cluster. For each receptor–ligand pair in each pairwise comparison between two cell types, this generated a null distribution. We obtained a P-value for the likelihood of cell-type specificity of a given receptor–ligand complex by t-test. We then prioritized the interactions that are highly enriched between cell types based on the number of significant pairs and selected anti-viral relevant ones.

## SUPPLEMENTAL INFORMATION

is linked to the online version of the paper.

## ACKNOWLEDGEMENTS

This study was funded by grants from the National Key Research and Development Program of China (2019YFA0110200, 2018YFC1200100, 2018ZX10301403), the special project for COVID-19 of Guangzhou Regenerative Medicine and Health Guangdong Laboratory(2020GZR110106006), the emergency grants for prevention and control of SARS-CoV-2 of Ministry of Science and Technology (2020YFC0841400) and Guangdong province (2020B111108001, 2018B020207013). We thank the patient who took part in this study.

## AUTHOR CONTRIBUTIONS

J.C., J.Z., J.Z. and Y.L. conceived and supervised the study, X.L., A.Z., Z.C., Y.X., F. Y., L.L., S.C., L.W., J.Z., F.L., D.C., R.C., N.Z., collected clinical specimen and executed the experiments. J.H., L.L., H.F., B.C., Y.M., L.L., Z.Z., J.S., Y.W., Y.Z., X.W., X.Z., N.Z., Y.H., H.L., J-Y.W., J.W., X.X., X.C. did single cell sequencing and bioinformatics analysis. J.C., J.Z., J.W. and J.Z. wrote the manuscript.

---

**Table S2.**

|  | Patient 1 | Patient 2 | Patient 3 | Patient 4 | Patient 5 |
| --- | --- | --- | --- | --- | --- |
| severity | severe | severe | mild | mild | severe |
| Age | 40s | 50s | 20s | 50s | 40s |
| Gender | male | male | female | male | male |
| Wuhan exposure history | Yes | Yes | Yes | No | Yes |
| Symptom onset day | 2020.01.22 | 2020.01.20 | 2020.02.06 | 2020.02.02 | 2020.01.26 |
| Fever/Cough | Yes | Yes | Yes | Yes | Yes |
| Hospitalization data | 2020.01.28- Present | 2020.01.22-Present | 2020.02.08-2020.02.21 | 2020.02.02-2020.04.09 | 2020.01.29-2020.03.26 |
| Intensive Care | 2020.01.31-2020.03.09 | 2020.01.23-Present | No | No | 2020.02.15-2020.03.09 |
| Sputum sampling date (scRNA-seq) | 2020.02.14 | 2020.02.14 | 2020.02.14 | 2020.02.14 | No |
| Sputum sampling date (FACS) | No | No | 2020.02.20 | 2020.02.20 | 2020.02.20; 2020.03.03; 2020.03.24 |
| Prednisone (泼尼松) | Yes | No | No | No | Yes |
| Methylprednisolone (甲强龙) | Yes | Yes | No | No | No |
| Zadaxin (日达仙) | No | Yes | No | No | Yes |
| γ-Immunoglobulin (丙球) | No | Yes | No | No | Yes |

